# TFF1 – a new Biomarker in Liquid Biopsies of Retinoblastoma under Therapy

**DOI:** 10.1101/2023.08.23.23294357

**Authors:** Maike Anna Busch, André Haase, Emily Alefeld, Eva Biewald, Leyla Jabbarli, Nicole Dünker

## Abstract

Effective management of retinoblastoma (RB), the most prevalent childhood eye cancer, depends on reliable monitoring and diagnosis. A promising candidate in this context is the secreted trefoil family factor peptide 1 (TFF1), recently discovered as a promising new biomarker in patients with a more advanced subtype of retinoblastoma. The present study investigated TFF1 expression within aqueous humor (AH) of enucleated eyes and compared TFF1 levels in AH and corresponding blood serum samples from RB patients undergoing intravitreal chemotherapy (IVC). TFF1 was consistently detectable in AH, confirming its potential as a biomarker. Crucially, our data confirmed that TFF1 secreting cells within the tumor mass originate from RB tumor cells, not from surrounding stromal cells. IVC therapy responsive patients exhibited remarkably reduced TFF1 levels post-therapy. By contrast, RB patients’ blood serum displayed low to undetectable levels of TFF1 even after sample concentration and no therapy-dependent changes were observed. Our findings suggest that compared to blood serum AH represents the more reliable source for TFF1 if used for liquid biopsy RB marker analysis in RB patients. Thus, analysis of TFF1 in AH of RB patients potentially provides a minimal invasive tool for monitoring RB therapy efficacy, suggesting its importance for effective treatment regimens.

**Simple Summary:** Effective management of retinoblastoma (RB), a common childhood eye cancer, requires accurate diagnosis and monitoring during therapy. In this study the liquid biopsy marker potential of the secreted trefoil family factor peptide 1 (TFF1), described as a biomarker of a more advanced RB subtype, was explored. TFF1 expression levels were investigated in aqueous humor (AH) of RB patients after enucleation and in RB patients undergoing intravitreal chemotherapy and compared with TFF1 expression levels in RB patients’ blood serum. AH showed consistent TFF1 levels in a subgroup of RB patients, remarkably decreasing post-therapy in responsive patients. Blood serum of RB patients only displayed low to non-detectable and therapy-independent TFF1 levels. The study suggests TFF1 expression in AH as a reliable biomarker, aiding RB diagnosis and treatment assessment and highlights it’s potential for non-invasive RB therapy monitoring.

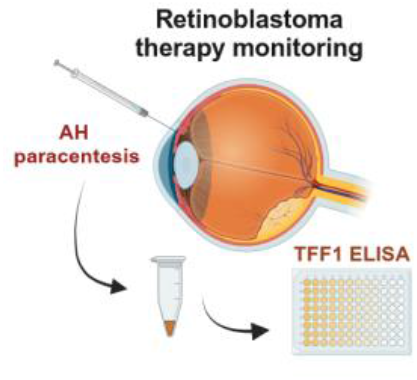

## 1. Introduction

Retinoblastoma (RB), the most common pediatric ocular malignancy, arises from the uncontrolled proliferation of developing retinal cells [1,2] and is characterized by the loss or mutation of both copies of the *RB1* gene, which regulates the cell cycle and inhibits tumorigenesis [3–6]. The disease manifests predominantly in children under five years of age, affecting both eyes in approximately 40% of all cases [5]. If left untreated, retinoblastoma can lead to severe visual impairment and, in most unfavorable cases, metastatic spread through the optic nerve into the central nervous system [1,7,8]. Traditional therapeutic methods for RB involve invasive procedures such as enucleation of the affected eye or systemic chemotherapy, which leads to lifetime visual limitations posing various risks to the patient [9–11]. Intravitreal chemotherapy (IVC) has emerged as a highly effective therapeutic modality for retinoblastoma, particularly for intraocular tumors [12–18], involving the direct injection of chemotherapeutic agents into the vitreous cavity of the affected eye, allowing for targeted treatment and reduced systemic toxicity. While IVC has shown promising results in managing RB, therapy monitoring remains crucial for the assessment of treatment efficacy and early detection of potential recurrence. Tissue biopsy is generally considered contraindicated for RB, as it is believed to promote extraocular spread [19]. Nevertheless, in some cases tissue biopsy is relevant as it allows for a reliable confirmation of RB diagnosis and the assessment of the *RB1* mutational status for prognostic counseling [20]. Despite the use of optical coherence tomography and B-scan ultrasonography, several differential diagnoses such as Coats disease, persistent fetal vasculature, retinopathy of prematurity, coloboma, and toxocariasis may be misdiagnosed as RB [19], eventually resulting in enucleation of infants for unequivocal diagnostic purposes [21].

Liquid biopsy offers a non-invasive alternative by detecting tumor-derived components, such as circulating tumor cells (CTCs), cell-free DNA (cfDNA), exosomes, microRNAs, and other secreted factors, in easily accessible body fluids like blood or aqueous humor (AH) [22–24]. Aqueous humor paracentesis is a straightforward and safe procedure commonly conducted under general anesthesia in conjunction with eye examinations in RB infants, which can also be combined with intravitreal administration of chemotherapy [24]. In recent decades, the identification of specific biomarkers - also detectable in in liquid biopsies - revolutionized cancer research and clinical practice [25–27]. Biomarkers, as measurable indicators of biological processes or disease states, play a crucial role in early detection, diagnosis, prognosis, and the development of targeted therapies for various malignancies [28,29] including RB. This technique not only presents an opportunity for early detection but also allows for real-time monitoring of disease progression and response to treatment [30,31].

Among the potential biomarkers for RB, trefoil factor family peptide 1 (TFF1) has emerged as an intriguing candidate with promising implications for cancer management [1,22,32]. TFF1, a member of the trefoil factor family peptides, plays crucial roles in maintaining mucosal integrity and promoting epithelial repair in various tissues [33–36]. Recent studies suggested a potential link between TFF1 and tumorigenesis, e.g clinico-pathological features, highlighting the diagnostic and prognostic value of TFF1 as a cancer biomarker. Previous studies by our group showed that RB cells lines and RB tumors express variable levels of TFF1 [46–48], while it is not expressed in the healthy human retina. Most recently we could demonstrate that TFF1 is also detectable in the aqueous humor of RB patients [23], rendering it a highly promising candidate as a RB biomarker in liquid biopsies. As a secreted, extracellular protein, TFF1 can be detected in body fluids [23,40,41], circumventing the need for invasive procedures. Besides a prospective biomarker like TFF1 potentially aids at early cancer detection, monitoring treatment response, and assessing disease progression, thereby improving patient outcomes.

In the study presented, we will shed light onto the potential of TFF1 as a non-invasive diagnostic and prognostic marker in RB liquid biopsies. TFF1 levels in RB patients’ aqueous humor and blood may not only enable early cancer detection, but also serve as a valuable indicator for monitoring therapy efficacy. Changes in TFF1 expression or release during the course of RB treatment might provide insights into treatment response and help to identify patients who may require additional interventions. Integrating TFF1 assessment into liquid biopsy protocols for RB patients could enhance therapeutic decision-making and improve long-term outcomes. As such, a comprehensive understanding of TFF1 as a cancer biomarker holds promise in shaping the future of RB diagnostics and personalized treatment approaches, ultimately contributing to improved clinical outcomes and quality of life for affected children.

## 2. Materials and Methods

### 2.1. Human retinoblastoma tumor, aqueous humor and blood samples

Human retinoblastoma (RB) primary tumor material, aqueous humor samples from enucleations of 8 patients as well as aqueous humor and blood serum samples from 7 RB patients under therapy and 6 healthy individuals (control group) were used for TFF1 expression studies. The Ethics Committee of the Medical Faculty of the University of Duisburg-Essen approved the use of retinoblastoma samples (approval # 14-5836-BO) for research conducted in the course of the study presented and written informed consent has been obtained from patients’ relatives or parents.

Primary tumor material and aqueous humor samples of eight patients were harvested immediately after enucleation (T27, T31, T32, T34, T36, T38, T40 and T41). Aqueous humor was aspirated via an anterior chamber puncture using a 30 G needle. In the next step, the actual tumor was removed from the globe via scleral fenestration. Aqueous humor and blood serum samples of seven patients were harvested under anesthesia prior to IVC treatment with melphalan. Subsequently, blood was centrifuged at 2,500 G for 15 min at 18°C. Separated serum fraction aliquots were stored at minus 80°C until further use. Besides, blood was drawn from six healthy individuals as a control group. Aqueous humor were stored at minus 80°C until use or further processing (see below), and tumor tissue samples were cultured as described below.

This study includes a case series of eight untreated eyes from individual children diagnosed with intraocular retinoblastoma (out of 2022) and seven treated eyes from individual children diagnosed with intraocular retinoblastoma between 2022 and 2023. Diagnosis of the untreated eyes was confirmed by a specialized pathologist after enucleation. The data collected included patient’s age at diagnosis, gender, laterality, ICRB stage (International Classification of Retinoblastoma), RB1 mutation status, tumor volume/size, optic nerve and choroid invasion.

### 2.2. Primary RB Cell Culture

The primary RB tumor material was initially cut up to small pieces with a sterile scalpel and subsequently washed three times in PBS with a centrifugation step in between (800 rpm for 2 minutes). After the last washing step the tumor material was cultivated in Dulbecco’s modified Eagle’s medium (DMEM; PAN-Biotech, Aidenbach, Germany) with 15% fetal calf serum (FCS; PAN-Biotech, Aidenbach, Germany), 100 U penicillin/ml and 100 µg streptomycin/ml (Invitrogen, Darmstadt, Germany), 4 mM L-glutamine (Sigma-Aldrich, München, Germany), 50 µM ß-mercaptoethanol (Carl-Roth, Karlsruhe, Germany) and 10 µg insulin/ml (Sigma-Aldrich, München, Germany) at 37°C, 10% CO2 and 95% humidity as described previously (Busch et al. 2015). The cells separated in culture into suspension (RB tumor cells) and adherent populations (RB derived stroma cells) and were subsequently cultured separately. Supernatants from both subcultures were harvested and residual cells were removed by centrifugation. Cell culture supernatants were kept at -20° C until usage.

### 2.3. Blood serum concentration and TFF1 ELISA analysis

Right before use, blood serum samples were concentrated up to 5-fold using protein concentrator column (3 kDa MWCO, Thermo Fischer Scientific, MA, USA) following the manufacturer’s instructions. One hundred microliters aqueous humor samples and concentrated blood serum from RB patients were analyzed using a human TFF1 ELISA kit (ab213833, abcam, Cambridge, UK) according to the manufacturer’s protocol. The standard curve included in the kit was used to determine the concentration of the samples analyzed.

### 2.4. Immunhistochemistry and immunfluorescence stainings

TFF1 immunostaining was performed with the Vectastin Elite ABC kit (Vector Laboratories, Burlingame, CA, USA) as previously described by our group [48]. The following antibody were used: TFF1 (1:200, abcam, Cambridge, UK, # ab92377). Images were acquired using an Aperio ScanScope AT2 (Leica, Wetzlar, Germany) slide scanner.

For immunofluorescence staining of TFF1, 1 x 10^5^ cells were seeded on poly-D-lysine (Sigma, Hamburg, Germany) coated coverslips and stained as previously described by our group [23]. Pictures were taken with a NIKON Eclipse E600 microscope equipped with a digital camera and NIKON Eclipse net software.

### 2.5. Statistical analysis

Statistical analyses were performed using GraphPad Prism 9. Results were analyzed by a Student’s t-test and considered significantly different if *p < 0.05, **p < 0.01, ***p < 0.001 or ****p<0.0001.

## 3. Results

### 3.1. Soluble TFF1 in aqueous humor of RB patients is secreted by RB tumor cells

In a most recent study we discovered TFF1 expression in a specific subgroup of retinoblastoma (RB) tumors with advanced stages, and found this soluble peptide to be secreted into the aqueous humor of RB patients [23]. To expand our investigation to a larger cohort of RB patients, we analyzed aqueous humor (AH) samples from eight additional RB patients after enucleation. Table 1 summarizes the clinical and pathological characteristics of the RB patients analyzed.

**Table 1.**
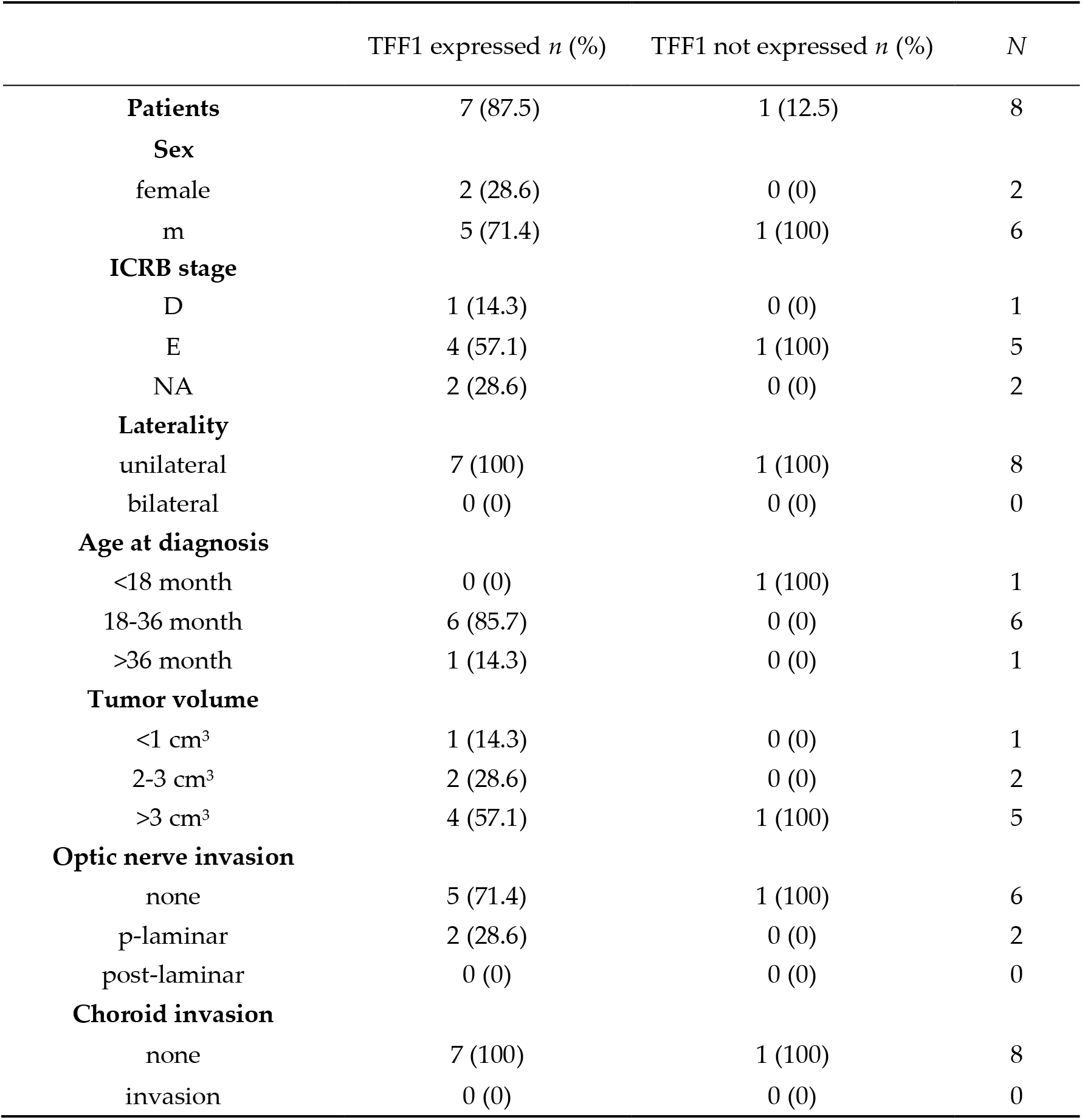
Clinical and pathological characteristics of RB patients stratified by TFF1 expression in aqueous humor. NA: not available, n: number in each group, N: total number, ICRB: International Classification of Retinoblastoma.

Using a specific, highly sensitive TFF1 ELISA (Figure 2a) allowed us to confirm that RB tumor cells secrete soluble TFF1 into the aqueous humor of RB patients’ eyes.

**Figure 1.**
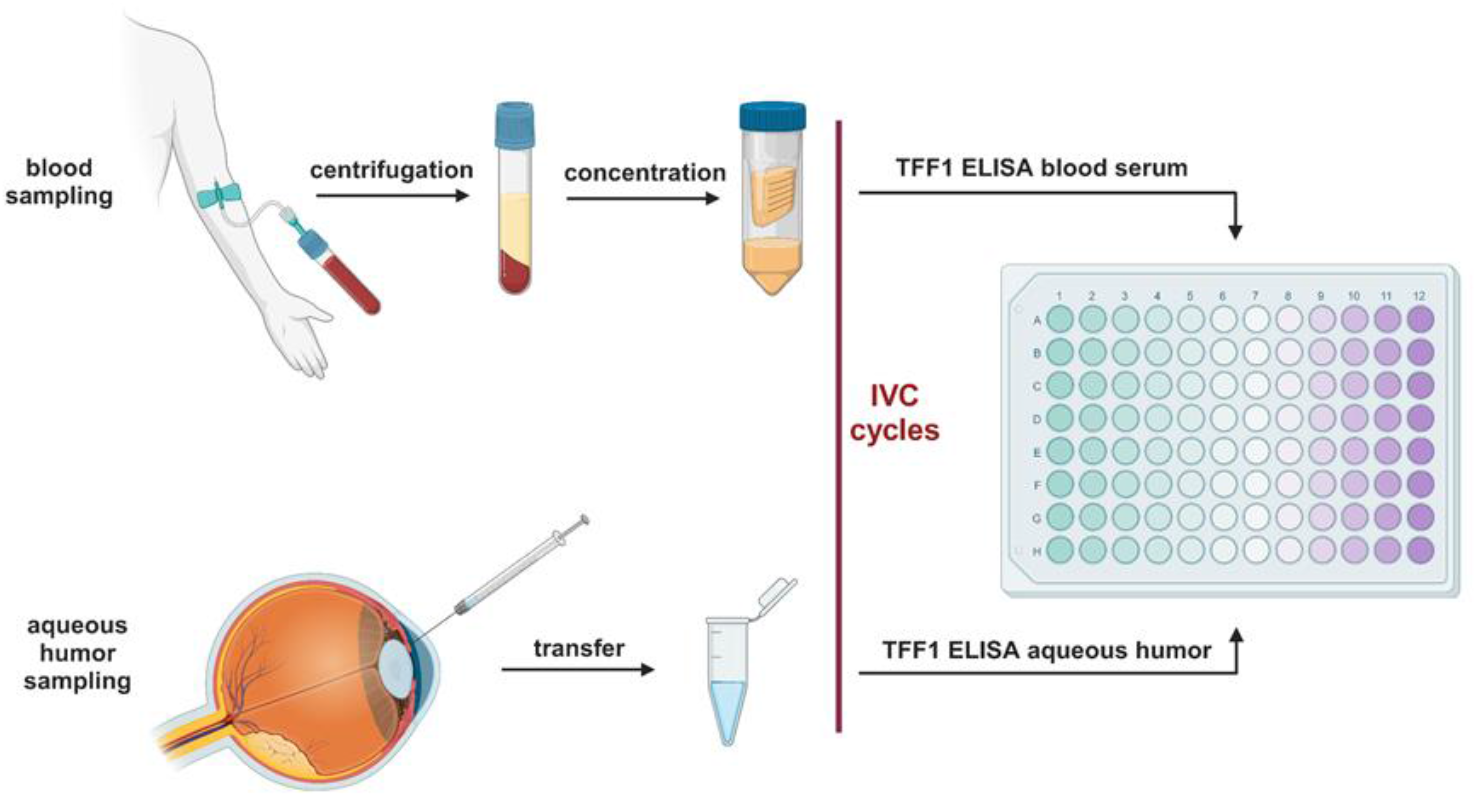
Workflow diagram of the TFF1 ELISA analysis. Created with BioRender.com.

**Figure 2.**
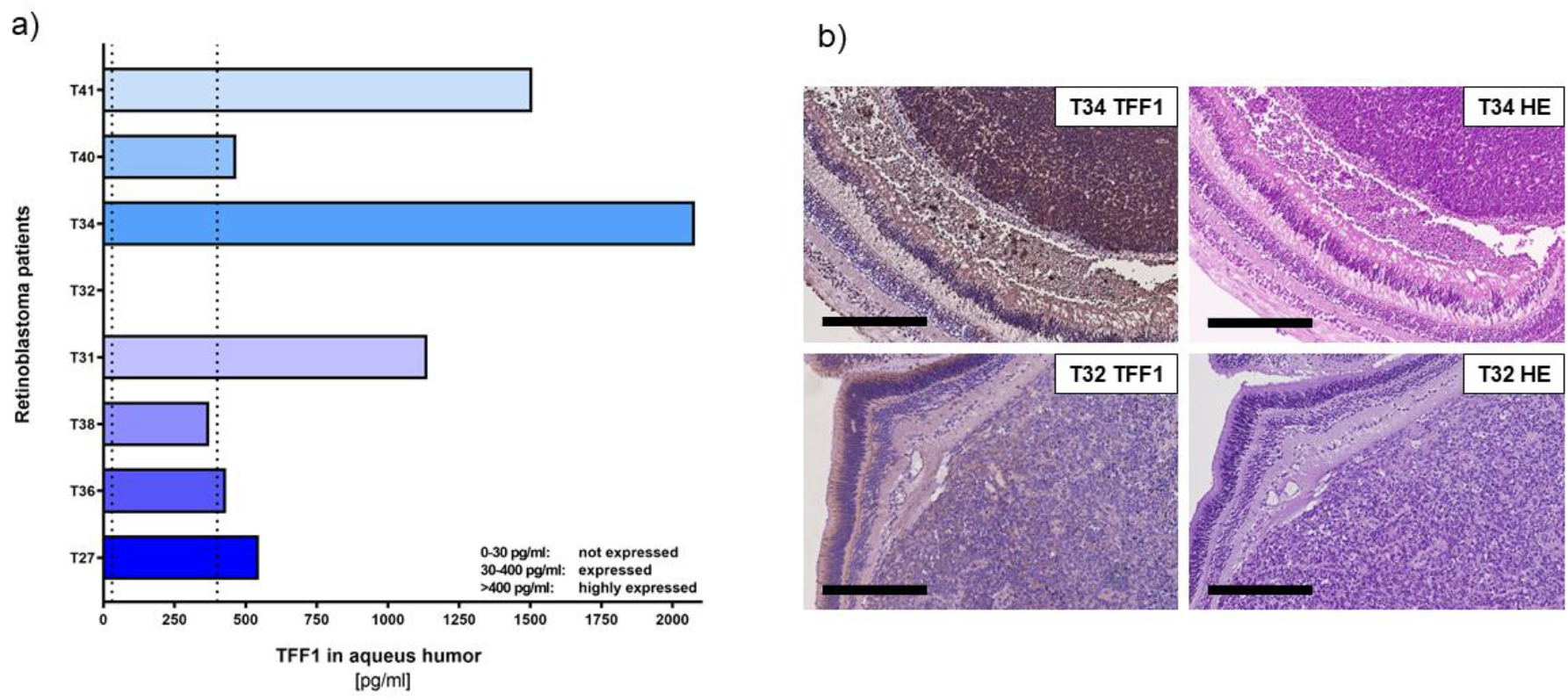
TFF1 expression analyses in aqueous humor samples and corresponding histological analysis of RB tumors in enucleated eyes of RB patients. (a) TFF1 ELISA analysis of 8 aqueous humor samples of RB patients, displaying six tumors highly expressing TFF1 (>400 pg/ml), one patient’s tumor with average TFF1 expression (30-400 pg/ml) and one patient without TFF1 expression (0-30 pg/ml) in the aqueous humor. Vertical dotted lines indicate three TFF1 expression levels. (b) TFF1 expression in the corresponding primary tumors is shown exemplarily for T34 (high TFF1 expression in AH) and T32 (no TFF1 expression in AH). Immunohistochemistry was revealed using diaminobenzidine detection (brown signal) and hematoxylin counterstaining (blue nuclei staining). Scale bars: 300µm.

Six out of eight tumors analyzed secreted high concentrations of TFF1 (ranging between 1000 and 4500 pg/ml; labeled as T27, T36, T31, T40, and T41) into the AH. Additionally, we found one tumor (T38) with moderate levels of TFF1 secretion. Only one (T32) out of eight tumors did not secrete any detectable TFF1 into the AH.

In order to compare the expression pattern of TFF1 in AH with its expression in original RB tumor specimens, we performed immunhistochemical staining for TFF1 on paraffin sections of enucleated patients eyes baring the investigated tumors. Remarkably, all tumors displaying detectable TFF1 in the AH samples also stained positive for TFF1 in the primary tumor sections (Figure 2b).

These results further support our previous findings and indicate that detection of TFF1 in RB patients’ AH represents a reliable marker for the presence of TFF1-secreting RB tumor cell. This finding holds promise for potential applications in RB patient monitoring and treatment strategies.

To investigate whether TFF1 is exclusively secreted by RB tumor cells and not by surrounding stromal tissue, we compared supernatants of a primary stromal cell culture and a primary RB tumor cell culture, both derived from enucleations of RB tumor baring patient eyes. The primary stromal cells did not carry the RB1 mutation present in the primary RB tumor cells, indicating their non-tumor identity. As expected, ELISA analysis revealed no detectable TFF1 secretion in the supernatant of the stromal cell culture (data not shown).

To further validate these results, we conducted immunofluorescence staining to assess the cellular expression of TFF1. The intracellular TFF1 expression pattern closely correlated with the TFF1 secretion status observed in the RB tumor cells and RB tumor-derived stromal cells. Specifically, primary RB tumor cells exhibited high levels of TFF1 expression (Figure 3a), while the corresponding RB derived stromal cells showed no detectable TFF1 expression (Figure 3b).

**Figure 3.**
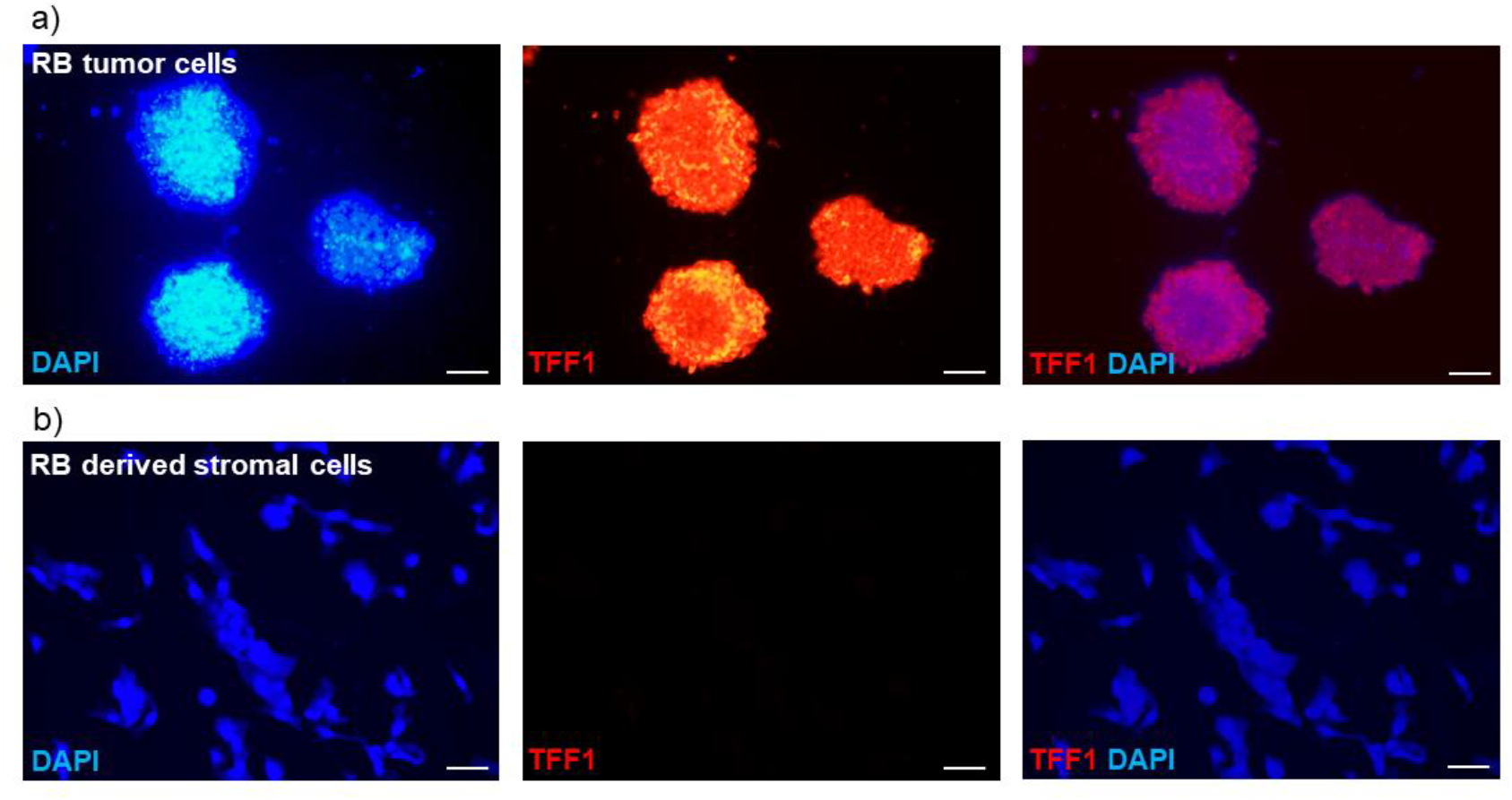
Immunofluorescence TFF1 staining of primary RB cell cultures. (a) Pictures of a primary cell culture of RB tumor cells and (b) corresponding RB tumor derived stromal cells in DAPI (blue), TFF1 (red) and merged DAPI/TFF1 immunofluorescenc staining (200x). RB tumor cells showed a high expression of TFF1 in contrast to the corresponding stromal cells, displaying no TFF1 expression. Scale bars: 50 µm

In summary, our previous findings [23] were confirmed by analyzing AH samples from enucleated RB eyes via TFF1 ELISA. Additionally, we demonstrated that the cells secreting TFF1 originate from tumorigenic cells and not from the stromal compartment of the RB tumor mass. This discovery highlights TFF1 as a potential marker for minimal invasive therapy monitoring via AH aspiration.

### 3.2. Analysis of soluble TFF1 secretion in AH and blood of RB patients under therapy

Monitoring and diagnosing RB is crucial to distinguish it from other diseases, evaluate treatment effectiveness, and identify potential recurrences. However, as RB tumor biopsies are not feasible, there is an urgent need for reliable biomarker to determine diagnosis and treatment success in non-enucleated RB tumors.

To investigate if secreted TFF1 expression changes in liquid biopsies during therapy, we examined a series of AH and corresponding blood samples from 7 RB patients using TFF1 ELISA. Liquid biopsies (AH and blood) were collected before the indicated intravitreal chemotherapy (IVC) treatment cycles with melphalan. We found that 3 out of the 7 RB patients (T28, T33, and T44) expressed soluble, secreted TFF1 in their AH (Figure 4a).

**Figure 4.**
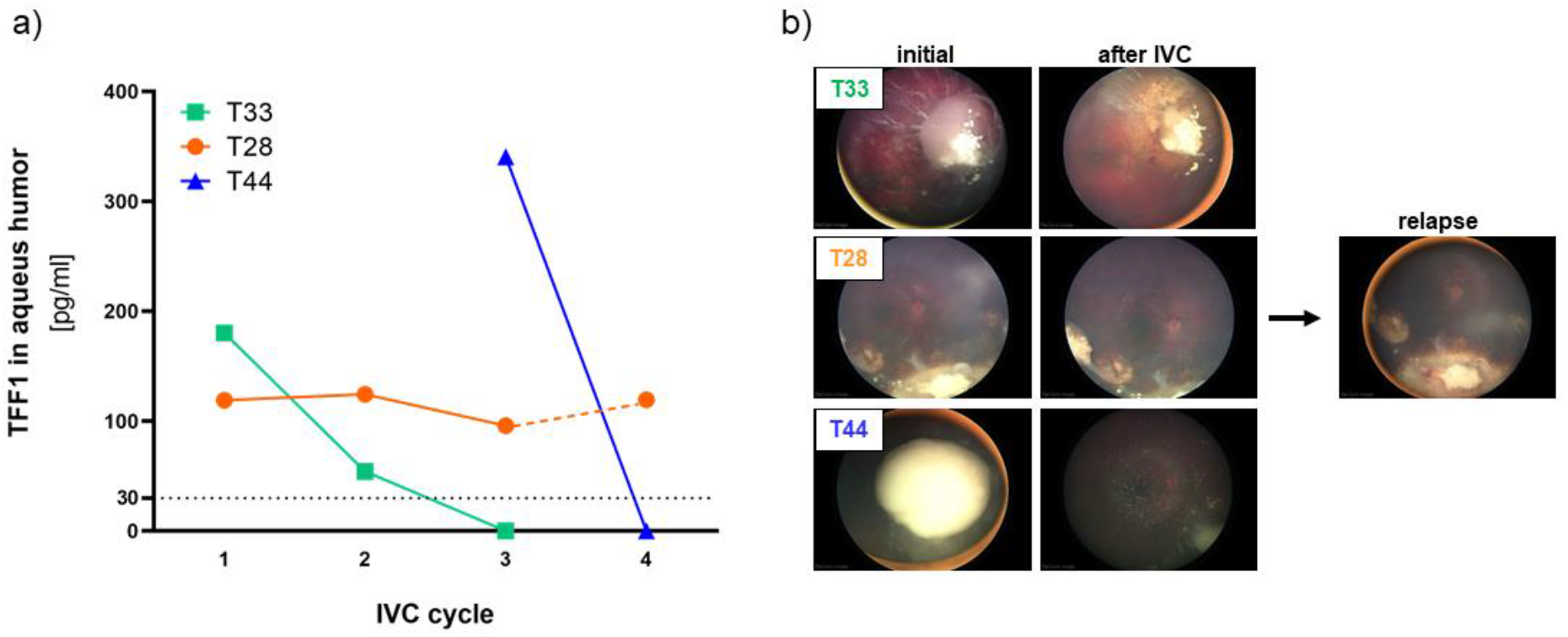
TFF1 expression analyses in aqueous humor samples of RB patients during therapy. (a) TFF1 ELISA analysis of three aqueous humor samples of RB patients taken prior to the indicated intravitreal chemotherapy (IVC) cycle with melphalan. All three patients initially expressed TFF1 (30-400 pg/ml) in the aqueous humor. Values below the vertical dotted line represent no TFF1 expression (0-30 pg/ml). (b) Fundoscopy pictures of of the three RB patients prior (initial) and after IVC with melphalan. For patient T28 a picture of the relapse after 5 month is provided.

The first AH sample of the tumor with the highest AH TFF1 concentration (T44) observed in our study was received from the clinics after two IVC cycles with melphalan. Thus, no information about the initial TFF1 concentration in the AH prior to therapy is available. We nevertheless included this specimen in our monitoring due to a remarkable decrease in TFF1 levels under therapy, which dropped to zero after only one additional IVC cycle.

Similarly, the tumor with the second-highest AH TFF1 concentration (T33) displayed a reduction to zero after only two therapy cycles. Interestingly, treatment outcome seems to correlate with the reduction of TFF1 expression in the AH in all RB tumors. Both tumors with TFF1 expression dropping to zero during IVC therapy responded well to treatment and showed positive outcomes as revealed by fundoscopy (Figure 4b) displaying a regression and calcification of the vitreous seeding. On the other hand, patient T28 exhibiting a constant TFF1 expression in the AH during therapy (Figure 4a, IVC cycle 1-3), exhibited a relapse after five month (Figure 4b), while still expressing high TFF1 levels in the AH (Figure 4a, IVC cycle 4).

These findings suggest that monitoring soluble TFF1 levels in the aqueous humor of RB patients under therapy could serve as a potential biomarker to assess treatment efficacy and predict therapeutic outcomes in non-enucleated RB tumors. This could significantly improve RB therapy management and patient care.

Additionally, we examined corresponding blood serum samples from the 7 RB patients, who underwent therapy and compared them to three control samples from non-RB children and three healthy adolescents. Detecting TFF1 in blood serum required prior concentration of the samples (as described in the materials and methods section) and even after concentration, only fairly low TFF1 levels (∼10 pg/ml) were detectable. Comparing TFF1 levels among individuals of the control groups we found young non-RB baring children under the age of six to display no detectable TFF1 expression compared to the healthy adolescent group (>14 years), expressing low levels of about 10 pg/ml TFF1 (Figure 5).

**Figure 5.**
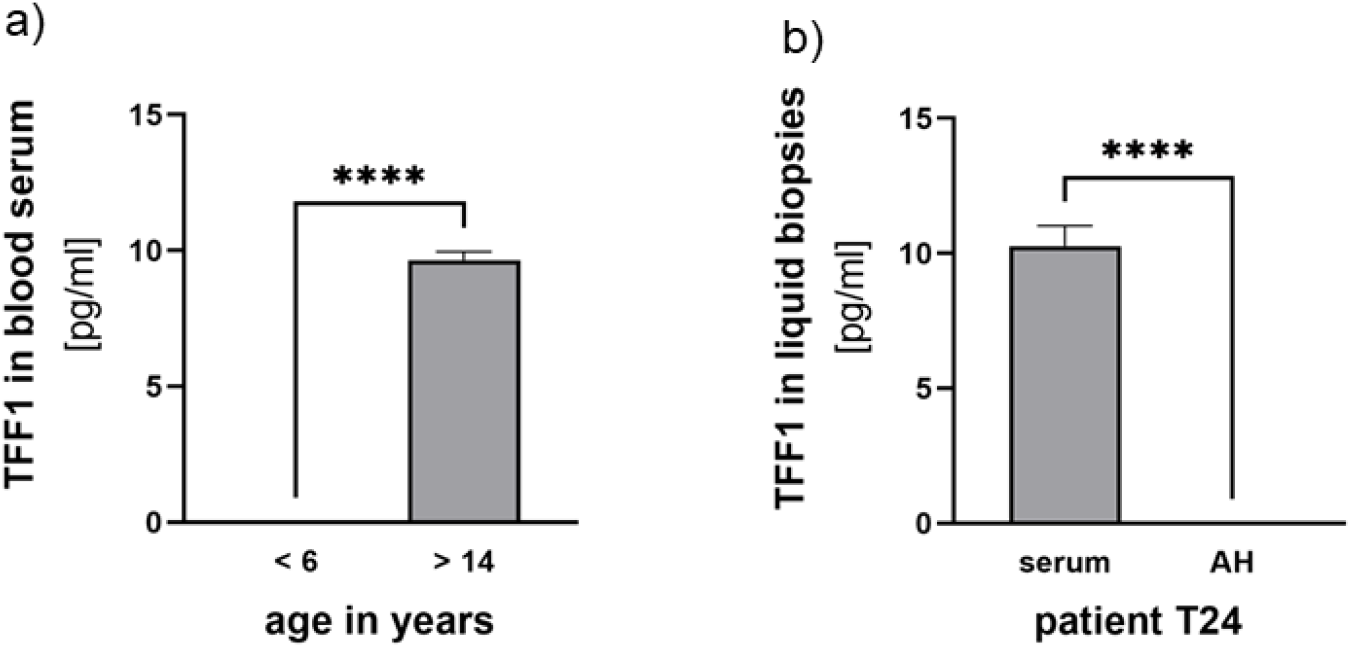
TFF1 expression analyses in blood serum and aqueous humor samples. (a) Comparison of TFF1 blood serum levels in control groups of non-RB children under the age of 6 years with healthy individuals over 14 years. (b) Comparison of TFF1 blood serum levels (serum) with TFF1 expression in aqueous humor (AH) of patient T24. Values are means of three samples ± SEM. ****p<0.0001 statistical differences compared to the control group calculated by students t-test.

Among the seven RB serum samples investigated, we detected low concentrations of TFF1 in three samples (< 12 pg/ml). However, in these patients, no TFF1 was detectable in the AH. Notably, one of the RB patients displaying detectable TFF1 expression in the blood was of higher age (Figure 5b), suggesting that serum expression might be related to the patient’s age.

In one RB patient, TFF1 expression was neither detectable in blood nor in AH (T26, Table 2). Furthermore, patient T28, whose TFF1 expression in AH remained unchanged during therapy, showed no detectable TFF1 expression in blood serum (Table 2). For patients T33 and T44, whose AH TFF1 expression dropped to zero under therapy, TFF1 expression was only randomly found in individual blood serum samples without a distinct expression pattern (Table 2).

Overall, no therapy-dependent changes in TFF1 expression were detectable in blood serum of any of the RB patients studied (Table 2), rendering blood serum unsuitable for TFF1 biomarker analyses. These findings indicate that AH remains the reliable source for monitoring TFF1 levels and assessing treatment responses in RB patients.

Table 2 summarizes clinical and pathological characteristics as well as TFF1 expression levels found in AH and blood serum samples of the 7 RB patients analyzed during therapy.

**Table 2.** Clinical and pathological characteristics of RB patients under IVC therapy with melphalan stratified by TFF1 expression in liquid biopsies (aqueous humor and blood serum). Red and blue labeling indicates the respective tumor analyzed in case of bilaterality

| Case<sup>1</sup> | Sex | Age at diagnosis (month) | Lateraliy | ICRB stage | Tumor size (mm) | Enucleation | Optic nerve invasion (MRI) | Choroid invasion (MRI) | RB1 mutation | Sample number | AH TFF1 (pg/ml) | Serum TFF1 (pg/ml) |
| --- | --- | --- | --- | --- | --- | --- | --- | --- | --- | --- | --- | --- |
| T24<sup>#</sup> | m | t: 18-36<br>r: >36 | b | ri: B<br>le: E | ri: 4.5<br>le: >10 | le: PE | no | no | yes | 1 | 0 | 9.73 |
|  |  |  |  |  |  |  |  |  |  | 2 | 0 | 9.97 |
|  |  |  |  |  |  |  |  |  |  | 3 | 0 | 11.12 |
| T25 | f | 18-36 | u | D | 8.7 | no | no | yes | no | 1 | 0 | 11.55 |
|  |  |  |  |  |  |  |  |  |  | 2 | 0 | 0 |
|  |  |  |  |  |  |  |  |  |  | 3 | 0 | 10.6 |
|  |  |  |  |  |  |  |  |  |  | 4 | 0 | 0 |
|  |  |  |  |  |  |  |  |  |  | 5 | 0 | 0 |
|  |  |  |  |  |  |  |  |  |  | 6 | 0 | 0 |
| T26 | m | <18 | b | D | 9 (both) | ri: no<br>le: SE | no | ri: no<br>le: yes | yes | 1 | 0 | 0 |
|  |  |  |  |  |  |  |  |  |  | 2 | 0 | 0 |
|  |  |  |  |  |  |  |  |  |  | 3 | 0 | 0 |
| T28<sup>#</sup> | f | t: 18-36<br>r: >36<br>r2: >36 | b | E | ri: 12<br>le: 20 | ri: no<br>le: PE | ri: no<br>le: yes | ri: no<br>le: yes | yes | 1 | 118.98 | 0 |
|  |  |  |  |  |  |  |  |  |  | 2 | 124.37 | 0 |
|  |  |  |  |  |  |  |  |  |  | 3 | 95.9 | 0 |
|  |  |  |  |  |  |  |  |  |  | 4 | 119.41 | 0 |
| T33 | m | 18-36 | u | D | n/a | no | n/a | n/a | n/a | 1 | 180.21 | 10.65 |
|  |  |  |  |  |  |  |  |  |  | 2 | 54.28 | 9.23 |
|  |  |  |  |  |  |  |  |  |  | 3 | 0 | 0 |
|  |  |  |  |  |  |  |  |  |  | 4 | 0 | 0 |
|  |  |  |  |  |  |  |  |  |  | 5 | 0 | 0 |
|  |  |  |  |  |  |  |  |  |  | 6 | 0 | 9.33 |
|  |  |  |  |  |  |  |  |  |  | 7 | 0 | 0 |
|  |  |  |  |  |  |  |  |  |  | 8 | 0 | 0 |
|  |  |  |  |  |  |  |  |  |  | 9 | 0 | 9.83 |
|  |  |  |  |  |  |  |  |  |  | 10 | 0 | 10.14 |
|  |  |  |  |  |  |  |  |  |  | 11 | 0 | 0 |
| T39 | f | 18-36 | b | ri: E<br>le: D | ri: 22<br>le: 7 | ri: SE<br>le: no | ri: yes<br>le: no | ri: yes<br>le: no | yes | 1 | 0 | 9.93 |
|  |  |  |  |  |  |  |  |  |  | 2 | 0 | 0 |
|  |  |  |  |  |  |  |  |  |  | 3 | 0 | 9.98 |
| T44 | m | >36 | u | C | 8 | no | no | no | n/a | 1<sup>*</sup> | 340.58 | 10.57 |
|  |  |  |  |  |  |  |  |  |  | 2 | 0 | 9.84 |

m: male, f: female, u: unilateral, b: bilateral, ri: right, le: left, t: primary tumor r: relapse, r2: second relapse, ICRB: International Classification of Retinoblastoma, SE: secondary enucleation, PE: primary enucleation, n/a: not available, #: relapse was analysed by TFF1 ELISA, \*: specimen after two cycles of melphalan, AH: aqueous humor, MRI: magnetic resonance imaging, <sup>1</sup>: case identifiers used are not known to anyone outside the research group and cannot reveal the identity of the study subjects

## 4. Discussion

In contrast to other cancer entities, molecular characterization of RB tumors mainly relays on tumor samples derived from enucleations, as direct tumor biopsies bare the risk of cancer cell seeding and spread outside the eye [49–51]. The identification of tumor biomarkers in RB liquid biopsies like aqueous humor and blood serum holds the potential to improve diagnosis and therapy management of this childhood eye cancer without the need for enucleation. Aqueous humor has been suggested as a surrogate for retinoblastoma tissue [25,52]. It can safely be aspirated from retinoblastoma eyes and contains tumor-derived cfDNA, proteins and metabolic targets, but also potential biomarkers like TFF1 [13,16,23,52–58]. Paracentesis of AH is minimally invasive and it is routinely aspirated from RB eyes undergoing salvage therapy with intravitreal injection of chemotherapeutics like melphalan. Nevertheless, this procedure entails the insertion of a fine-gauge needle through the cornea, which carries a minimal risk of complications such as bleeding, infection, cataract formation, iris trauma, and in theory, the potential for tumor cells to spread to the orbit [59]. This raises the question if blood, a less invasive sample to obtain, could also be a source for RB biomarkers like TFF1.

Our study presented aimed to explore the potential of TFF1 as a biomarker for retinoblastoma in aqueous humor and blood serum of patients in general and under therapy. Monitoring liquid biopsies biomarker levels in RB diagnosis and under therapy may ultimately enable a timely correlation between TFF1 expression levels in aqueous humor and/or blood serum and RB progression. TFF1 has previously been identified as a functional biomarker in various other types of tumors, such as breast cancer [44,60], esophageal squamous cell carcinoma [61], and gastric cancer [62]. Notably, in breast cancer a correlation between elevated TFF1 expression in blood samples of patients with compared to those without metastatic disease was observed [42]. By evaluating TFF1 staining in tumor sections post-enucleation, we and others already suggested TFF1 as a potential biomarker for a specific subset of retinoblastomas [1,32]. We could initially demonstrate that TFF1 correlates with a higher clinical tumor-node-metastasis (TNM) stage and poorly differentiated tumor cells [32], later identified and specified as RB subtype 2 with a higher risk of metastasis by Liu et al [1]. Most recently, we revealed for the first time that soluble TFF1 is secreted into the AH of RB patients [23].

Here we analyzed AH of eight patients after enucleation as well as AH and corresponding blood serum of seven RB patients under therapy for TFF1 expression and secretion status. In addition, we investigated TFF1 expression in blood serum samples of control specimens including young children and adolescents in order to compare TFF1 levels of both groups.

The study presented verifies our previous findings [23] that TFF1 is secreted into the AH of most patients analyzed after enucleation. All patients with TFF1 positive AH also expressed TFF1 in the original tumor. Furthermore, we investigated if TFF1 is secreted exclusively by RB tumor cells or also by tumor associated stromal cells. We therefore analyzed primary RB tumor cells and compared their endogenous TFF1 expression status and ability to secrete TFF1 into the supernatant with RB tumor derived stromal cells. We could show that only RB tumor cells and not RB derived stromal cells express and secrete TFF1, rendering TFF1 a specific marker for RB tumor cells.

Analyzing AH of RB patients under therapy, we found three out of seven patients initially secreting soluble TFF1 into the AH. Two of these patients completely lost TFF1 expression in their AH during IVC therapy with melphalan, indicating a direct influence of the therapy regimen on TFF1 expression. The most obvious mechanistic explanation for this loss could be the death of TFF1 expressing RB tumor cells during therapy suggesting TFF1 a direct monitor for residual living tumor cells. In line with these results, one RB tumor investigated displayed unvaried high TFF1 secretion into the AH during therapy and developed a relapse after five month possibly indicating persisting RB tumor cells not readily visible in funduscopy and sonography. One might hypothesize that particularly tumors with a high stromal content respond well to therapy, but it may be mainly stromal and not tumor cells which are susceptible to the treatment, a distinction that cannot be made by funduscopy. In this case, TFF1 could be an extraordinary helpful marker for monitoring residual RB tumor cells, still secreting this biomarker indicating the need for higher frequent post-therapy screening of the respective RB patients.

Furthermore, we investigated if TFF1 can also be detected in blood serum of RB patients and if TFF1 levels might correlate with therapy efficacy. Some RB patients’ blood samples displayed detectable, yet very low concentrations of soluble TFF1, however without any correlation to TFF1 levels in the corresponding AH samples and with no correlation to the treatment outcome. Even in the control groups, only very low concentrations of TFF1 were detectable. Thus, AH seems to be superior to blood serum as a liquid biopsy for RB not only for detecting tumor-associated chromosomal changes in whole genome sequencing [59], but also for TFF1 monitoring. Interestingly, however, no TFF1 expression was detectable in non-RB children under the age of six in comparison to the older, adolescent control group (> 13 years), displaying low, but traceable levels. This may lead to the assumption that healthy young children do not secrete TFF1 into the blood.

Summarizing one can state that the analysis of TFF1 in the AH of RB patients opens the field for additional diagnostic approaches and therapy monitoring, using TFF1 as a potential biomarker for RB tumor cells (Figure 6).

**Figure 6.**
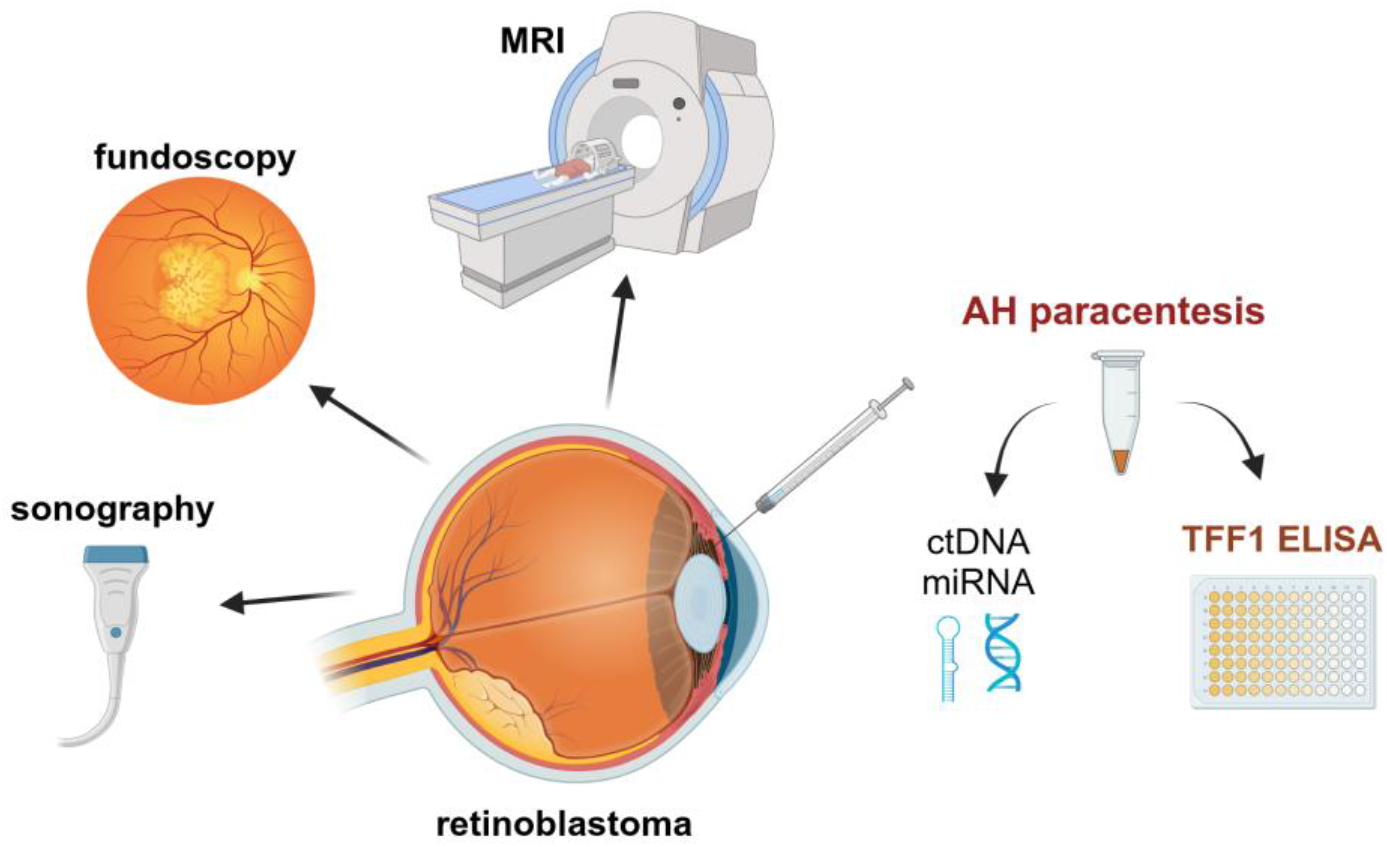
Potential monitoring procedures of RB patients under treatment to improve the outcome. Created with ©BioRender.com.

Nevertheless, it is necessary to further investigate TFF1 secretion into the AH during therapy to develop a save future implementation of TFF1 in clinical diagnostic and treatment regimens.

## 5. Conclusions

Analyzing liquid biopsies and identifying tumor biomarkers hold the potential to improve the management of retinoblastoma therapy and provide clarity in clinically uncertain differential diagnoses. TFF1 is supposed to be a marker of a subset of more advanced RB tumors and its expression correlates with a higher risk for metastases. We provided evidence for TFF1 therapy dependent expression changes in the AH of RB patients, strongly suggesting TFF1 as a clinically interesting new RB biomarker in therapy monitoring.

## Data Availability

All data produced in the present work are contained in the manuscript

## Author Contributions

Conceptualization, MAB; methodology, AH and EA; validation, MAB and AH; formal analysis, AH and EA; investigation, AH and EA; resources, EB and LJ; data curation, AH and MAB; writing—original draft preparation, MAB and ND; writing—review and editing, MAB and ND; visualization, MAB and AH; supervision, MAB and ND; project administration, MAB and ND; All authors have read and agreed to the published version of the manuscript.

## Institutional Review Board Statement

The study was conducted according to the guidelines of the Declaration of Helsinki, and approved by the Ethics Committee of the Medical Faculty of the University of Duisburg-Essen (approval # 06-30214; date of approval: 5/12/2006; approval # 14-5836-BO; date of approval: 11/3/2020).

## Informed Consent Statement

Informed consent was obtained from all subjects involved in the study.

## Acknowledgments

The authors would like to thank N. Bechrakis and T. Kiefer for their valuable support and A. Bollmeier for excellent technical assistance.

